# Evaluation of the accuracy and ease-of-use of Abbott PanBio - A WHO emergency use listed, rapid, antigen-detecting point-of-care diagnostic test for *SARS-CoV-2*

**DOI:** 10.1101/2020.11.27.20239699

**Authors:** L.J. Krüger, M. Gaeddert, F. Tobian, F. Lainati, C. Gottschalk, J.A.F. Klein, P. Schnitzler, H.G. Kräusslich, O. Nikolai, A.K. Lindner, F.P. Mockenhaupt, J. Seybold, V.M. Corman, C. Drosten, N.R. Pollock, B. Knorr, A. Welker, M. de Vos, J.A. Sacks, C.M. Denkinger, for the study team

**Author notes:** To whom correspondence should be addressed: Claudia M. Denkinger. Members of the study team are detailed at the end of the paper.

## Abstract

**Background:** Diagnostics are essential for controlling the pandemic. Identifying a reliable and fast diagnostic is needed to support testing. We assessed performance and ease-of-use of the Abbott PanBio antigen-detecting rapid diagnostic test (Ag-RDT).

**Methods:** This prospective, multi-centre diagnostic accuracy study enrolled at two sites in Germany. Following routine testing with RT-PCR, a second study-exclusive swab was performed for Ag-RDT testing. Routine swabs were nasopharyngeal (NP) or combined NP/oropharyngeal (OP) whereas the study-exclusive swabs were NP. To evaluate performance, sensitivity and specificity were assessed overall and in predefined sub analyses accordingly to cycle-threshold values, days of symptoms, disease severity and study site. Additionally, an ease-of-use assessment and System Usability Scale (SUS) were performed.

**Findings:** 1108 participants were enrolled between Sept 28 and Oct 30, 2020. Of these, 106 (9∙6%) were PCR-positive. The Abbott PanBio detected 92/106 PCR-positive participants with a sensitivity of 86∙8% (95% CI: 79∙0% - 92∙0%) and a specificity of 99∙9% (95% CI: 99∙4%-100%). The sub analyses indicated that sensitivity was 95∙8% in CT-values <25 and within the first seven days from symptom onset. The test was characterized as easy to use (SUS: 86/100) and considered suitable for point-of-care settings.

**Interpretation:** The Abbott PanBio Ag-RDT performs well for *SARS-CoV-2* testing in this large manufacturer independent study, confirming its WHO recommendation for Emergency Use in settings with limited resources.

**Funding:** The Foundation of Innovative New Diagnostics supplied the test kits for the study. The internal funds from the Heidelberg University as well as the Charité Berlin supported this study.

## Introduction

Diagnostics are a corner-stone of pandemic control. The World Health Organisation (WHO) emphasized already in March, 2020 the importance of access to testing for the effective control of *SARS-CoV-2*.^1^ While reverse-transcriptase polymerase chain reaction (RT-PCR) remains the gold standard among all diagnostic tests for *SARS-CoV-2*, access may be limited due to shortages of instruments, supplies and experienced operators, particularly in resource-limited settings. Antigen-detecting tests offer an alternative to RT-PCR and have been recommended by the WHO for appropriate settings where nucleic acid amplification technology (NAAT) testing is limited or where prolonged turnaround times slow down clinical testing.^2^ The recommendation outlines that only Ag-RDT meeting the minimum performance requirements of ≥80% sensitivity and ≥97% specificity compared to a NAAT reference assay should be considered for use.^2^. Before implementation of Ag-RDTs a manufacturer-independent evaluation should be conducted to assess claims made by the manufacturer.

The Foundation of New Innovative Diagnostics (FIND) has identified 84 antigen tests in the pipeline for *SARS-CoV-2*.^3^ To this date, the WHO has recommended two Ag-RDTs on the Emergency Use Listing based on data from manufacturers as well as one independently-conducted accuracy study.^4^ These two Ag-RDT are SD Biosensor STANDARD Q (recommended on Sept 22, 2020) followed by the Abbott PanBio Ag-RDT (Oct 2,2020).^5,6^

Following the WHO recommendations, several studies evaluating the Abbott PanBio Ag-RDT have been published. Of the three large studies available to date only one single-centre study prospectively enrolled participants.^7^ The other two selected stored samples with the representativeness of the selection being unclear in one study. ^8,9^ The sensitivity was determined to be in the range of 82-92% in the three studies along with an excellent specificity of 98.9% and above.^7-10^ To be able to generate additional independent data the WHO called on Nov 10, 2020, for *SARS-CoV-2* antigen detecting rapid diagnostic test implementation projects.^11^

The prospective multi-centre clinical accuracy study reported here represents the largest manufacturer-independent dataset for the POC performance of the Abbott PanBio Ag-RDT o date and also represents to our knowledge the only comprehensive ease-of-use assessment of the Abbott PanBio Ag-RDT.

## Methods

### Ethic statement

The study protocol was approved in March 2020 by the ethical review committee at the Heidelberg University Hospital for the two study sites Heidelberg and Berlin in Germany (Registration number S-180/2020).

### Role of funding

The Foundation for Innovative New Diagnostics (FIND), WHO collaborating centre for Coronavirus disease 2019 (COVID-19) diagnostics, supplied the test kits for the study. Internal funds from the Heidelberg University as well as the Charité Berlin supported this study. FIND provided input on study design and data analysis in the form of an academic exchange with the rest of the study group. All data was at all times fully accessible to the corresponding author who made the final decision to submit this manuscript for publication.

### Clinical diagnostic accuracy

*The standards for reporting diagnostic accuracy studies* (STARD) was used for the transparency and completeness of this diagnostic accuracy study.

#### Test evaluated

The test evaluated in this clinical diagnostic accuracy study is the PanBio™ COVID-19 Ag Rapid Test Device^12^ (Abbott Rapid Diagnostics, Jena, Germany; henceforth called PanBio).

The test uses the lateral flow assay principle in a cassette format design for the detection of viral antigens. The test kits include proprietary swabs for sample collection. As indicated in the instruction for use (IFU), five drops of the extracted specimen in the provided buffer solution are applied to the test device. Colloidal gold conjugated antibodies on the membrane strip react with viral antigens and capture antibodies to generate a colour change in the device window, which can be interpreted with the naked eye. The results are interpreted between 15 minutes and 20 minutes of incubation and are considered invalid if interpreted after this timeframe. The manufacturer’s instructions for use (IFU) were followed during sampling and testing procedures.

#### Study design and participants

The enrolment of participants was conducted at two sites; Heidelberg and Berlin, Germany. In Heidelberg, participants presented at a drive-in testing site. In Berlin, participants were enrolled at a clinical ambulatory testing facility. Inclusion criteria for the participation were age ≥18 years and classification as being at risk for a *SARS-CoV-2* infection by the local health department based on proven contact with a confirmed *SARS-CoV-2* case or having suggestive symptoms for infection. Individuals with a prior positive RT-PCR test for *SARS-CoV-2* or those who could not give written informed consent due to limited command in English or German were excluded from enrolment. A protocol is available upon request.

#### Study procedures

Individuals presenting for routine testing and meeting the inclusion criteria were invited to participate in the study. After providing written informed consent, participants first underwent the routine swab directly followed by the study-exclusive swab for Ag-RDT testing, performed by the trained study team. Sampling for RT-PCR testing was performed with a nasopharyngeal (NP) swab in Heidelberg and a combination of a NP and oropharyngeal (OP) swab in Berlin as per institutional procedure. The sampling for the Ag-RDT PanBio was an NP swab; however, if NP swabbing was contraindicated for clinical reasons (e.g. risk of bleeding) an OP swab was performed. The second study-exclusive swab was taken in the same nostril as the routine swab, only if the participant indicated the preference for the other nostril, the swabbing was performed on the opposite site. Laboratory personnel working in both the Ag-RDT testing team and the RT-PCR laboratory were blinded to the results of the other test at all times.

#### Antigen-detecting testing

Ag-RDT testing was performed in immediate proximity to the sampling in a separate room/container. The laboratories were designed and the laboratory personnel were trained to prevent cross-contamination with areas predefined for handling either infectious or non-infectious material following detailed working procedures. The Ag-RDT test was started directly after sample collection. The test was conducted as indicated in the IFU, interpreting the test with the naked eye after 15 minutes by two readers blinded to the results of the other. In the case of discrepant results both readers re-interpreted the results and agreed on a final result. Invalid test results were repeated once with the remaining buffer solution in the test tubes.

#### RT-PCR testing

The collected swabs (Heidelberg, IMPROSWAB, Improve; Berlin, eSwab, Copan) for RT-PCR testing, kept in provided Amies solution, were processed in the referral laboratory following the established laboratory routines. The RT-PCR assays which were used as reference standards in the laboratories were the Allplex *SARS-CoV-2* Assay from Seegene (Seoul, South Korea) in Heidelberg and the Roche Cobas SARS CoV-2 assay (Pleasanton, CA United States) on the Cobas® 6800 or 8800 system or the SARS CoV-2 assay from TibMolbiol (Berlin, Germany) in Berlin. The RT-PCR was considered positive with CT-values below a predefined threshold as per manufacturer instructions. CT values varied in a range of 2-3 between the three technologies. For the CT-value presentation and the viral load calculations the E-Gene was used as reference CT-value. A conversion of the CT-values into viral-load was performed using quantified specific in vitro-transcribed RNA.^13^

#### Additional data collection

All participants were asked to provide additional information about their comorbidities, symptoms, symptom duration and severity of disease (questionnaire available in the supplement material, Section (B)). In Heidelberg, the participants were contacted via telephone or E-mail after leaving the drive-In testing site, depending on their indicated preference during enrolment on-site, to complete the questionnaire either with an interviewer or by themselves. In Berlin, the participant was asked to complete the questionnaire directly on-site before the completion of the sample collection.

### Data management

All data were collected and managed using Research Electronic Data Capture (REDCap) tools hosted at Heidelberg University. ^14,15^

### System Usability Scale and Ease-of-Use Assessment

A standardized System Usability Scale (SUS) questionnaire and an ease-of-use assessment (EoU) were designed to understand the usability and feasibility of the test.^16^ The questionnaire and the EoU survey can be found in the supplement material Section (C) and (D). Laboratory personnel from both study sites were invited to complete the questionnaire. An over-all SUS score above 68 is interpreted as above average and anything below the score of 68 is below average.^16^ A heat-map was generated to analyse aspects related to the ease-of-use of the test, categorising each as satisfactory, average or dissatisfactory. The matrix used for this analysis is also found in the supplement material Section (C).

### Statistical Analysis

The sensitivity and specificity of the Ag-RDT with 95% confidence intervals (95% CIs) were assessed as per Altman compared to RT-PCR as reference standard (statistical analysis plan available upon request).^17^ Sub analyses were performed by sampling strategy, symptoms, duration of symptoms, CT-values and study sites. The significance threshold was set at a two-sided alpha value of 0∙05. Participants with an invalid PCR result were excluded from the analysis. All analyses and plots were performed using R version 4.0.3.

## Results

### Clinical diagnostic accuracy

During the enrolment period, from Sept 28, 2020 to Oct 30, 2020, a total of 1261 eligible participants meeting the inclusion criteria were screened for this study. From these 1261 participants, 1119 agreed to undergo a second swab for study purposes only (Figure 1. Study Flow). 10 participants had to be excluded from the study, initially agreeing on participation but denying a second sample collection after the routine swab was performed. After the data cleaning and the exclusion of one invalid PCR test result (N=1), a total of 1108 participants were included in the analysis. The site in Heidelberg enrolled 858 participants between the Sept 28 and Oct 30, 2020, and the site in Berlin enrolled 250 participants between Oct 19-30, 2020.

**Figure 1:**
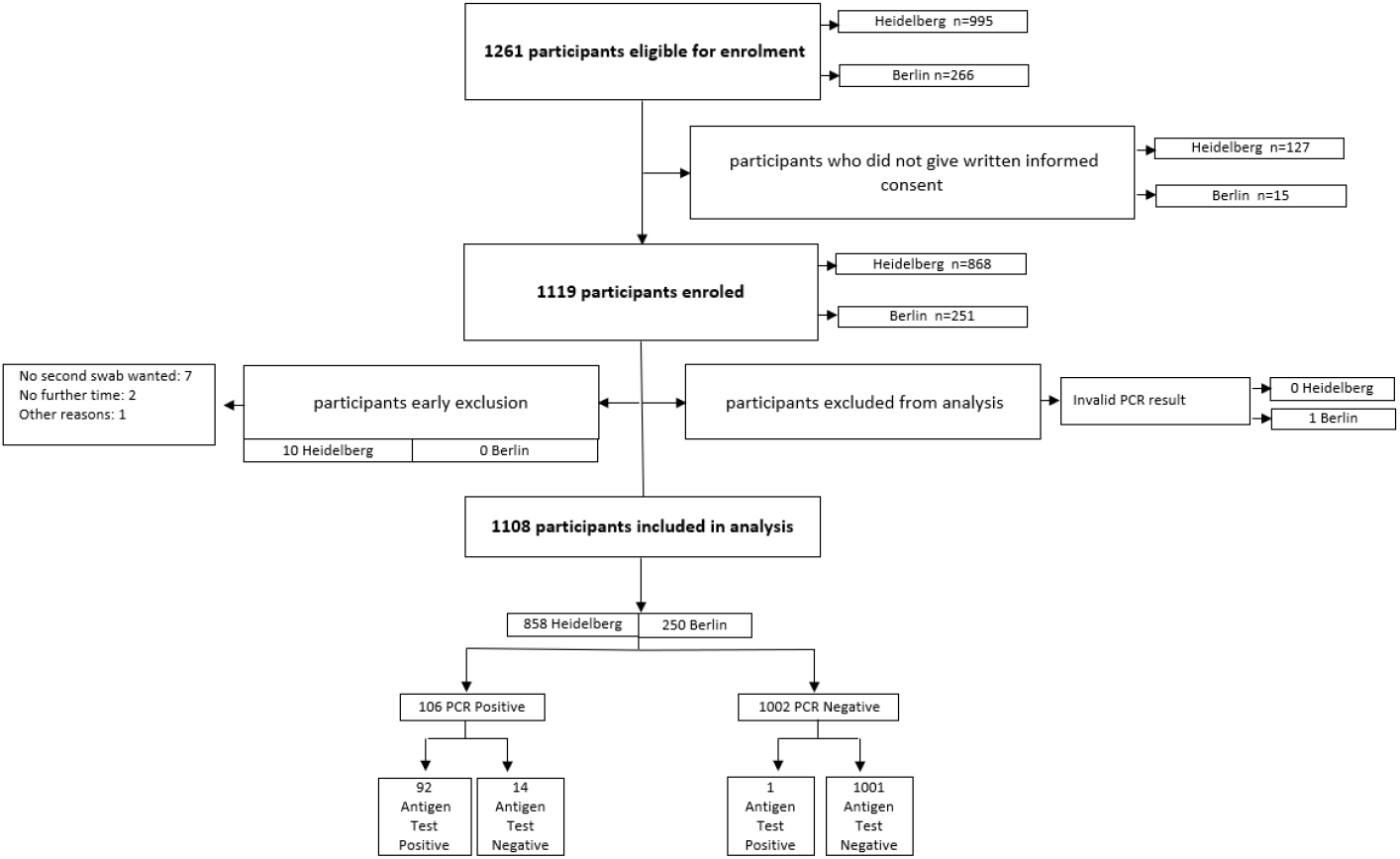
Study Flow.

The clinical and demographic characteristics of the enrolled participants are summarized in Table 1. The mean age of all participants was 39∙4 years (Standard Deviation (SD) 14∙1) with Berlin presenting a younger study population compared to Heidelberg. 50∙7% of participants were female and 33∙4% had comorbidities. 712 participants (64∙7%) reported having symptoms on the testing day, with an average symptom duration in days of 4∙01 days (SD 3∙1). The populations enrolled in Berlin and Heidelberg were significantly different in that participants in Berlin typically enrolled with symptoms (96∙8%) while in Heidelberg almost half of the participants were tested based on high risk contacts without symptoms and compared to the other half of participants reporting symptoms on the testing day (54∙8%). Also, participants in Heidelberg were more likely to present earlier in their course of disease (mean 3∙5 versus 4∙98 days in Berlin) and were more likely to have comorbidities (36∙6% in Heidelberg versus 21∙2% in Berlin). In total 106 (9∙6%) participants were diagnosed with a SARS-CoV-2 infection by RT-PCR testing during the enrolment period with 23∙6% in Berlin and 5∙5% in Heidelberg. The mean viral load was 7∙4 for both sites with only a slight difference in the SD (Table 1).

**Table 1:** Study population characteristics.

|  |  | Overall<br>Total N= 1108 | Heidelberg<br>Total N= 858 | Berlin<br>Total N= 250 |
| --- | --- | --- | --- | --- |
| **Age in years**<br>Information available on N=1108 | Mean<br>(SD) | 39.4<br>(14.1) | 40.3<br>(14.6) | 34.7<br>(11.5) |
| **Gender**<br>Information available on N=1099 | Female<br>N (%) | 557<br>(50.7%) | 455<br>(53.0%) | 102<br>(40.8%) |
| **BMI >25**<br>Information available on<br>N=1030 | N<br>(%) | 487<br>(47.3%) | 424<br>(49.4%) | 63<br>(24.8%) |
| **Comorbidities**<br>Information available on<br>N=1100 | N<br>(%) | 367<br>(33.4%) | 314<br>(36.6%) | 53<br>(21.2%) |
| **Symptoms on Testing Day<sup>#</sup>**<br>Information available on N= 1100 | N<br>(%) | 712<br>(64.7%) | 470<br>(54.8%) | 242<br>(96.8%) |
| **Duration of symptoms from day of testing**<br>**in days**<br>Information available on N=687 | Mean<br>(SD) | 4.01<br>(3.1) | 3.50<br>(2.83) | 4.98<br>(3.32) |
| **Previous test negative**<br>Information available on N= 885 | Yes<br>N (%) | 250<br>(28.2%) | 217<br>(25.3%) | 33<br>(13.2%) |
| **RT-PCR positives\*** | N<br>(%) | 106<br>(9.6%) | 47<br>(5.5%) | 59<br>(23.6%) |
| **Viral Load**<br>(log10 RNA SARS-CoV2/ml)<br>Information available on N=105 | Mean<br>(SD) | 7.4<br>(1.4) | 7.4<br>(1.3) | 7.4<br>(1.5) |

\*For one positive RT-PCR case CT-Value/Viral Load not provided due to a mutation in the E-Gene

The PanBio had an overall sensitivity of 86∙8% (92/106 RT-PCR positives detected; 95% Confidence Interval (CI): 79∙0% - 92∙0%) and a specificity of 99∙9% (1 false positive; 95% CI: 99∙4%-100%). In a predefined sub analysis by CT-value, sensitivity for samples that had a CT-Value >=25 was 66∙7% (95% CI: 49∙6%-80∙2%) and sensitivity for samples with a CT-value <25 was 95∙8% (95% CI: 88∙5%-98∙6%). When samples with a CT-value >=30 were assessed, the sensitivity was only 33% (95% CI 13∙8%-60∙1%) but 93∙5% (95% CI 86∙6%-97∙0%) for samples with a CT-value <30 (see Table 2 and Figure 2). The sensitivity decreased as viral load decreased (Figure 2). A detailed table summarizing the viral load equivalents to the CT-values for both study sites and by RT-PCR reference standard is provided in the supplement material Section (G) Table 3.

**Table 2:** Subgroup Analyses for PanBio.

|  | Overall | Ag-Test positive/<br>PCR positive | Ag-Test negative/<br>PCR positive | Ag-Test positive/<br>PCR negative | Ag-Test negative/<br>PCR negative | Sensitivity<br>(%, 95% CI) | Specificity<br>(%, 95% CI) |
| --- | --- | --- | --- | --- | --- | --- | --- |
| **Sensitivity** |  |  |  |  |  |  |  |
| **Overall** | 1108<br>(100%) | 92<br>(8.3%) | 14<br>(1.3%) | 1<br>(0.1%) | 1001<br>(90.3%) | 86.8%<br>(79.0%-92.0%) | 99.9%<br>(99.4%-100%) |
| **Heidelberg** | 858<br>(77.4%) | 44<br>(5.1%) | 3<br>(0.3%) | 0 | 811<br>(94.5%) | 93.6%<br>(82.8%-97.8%) | 100%<br>(99.5%-100%) |
| **Berlin** | 250<br>(22.6%) | 48<br>(19.2%) | 11<br>(4.4%) | 1<br>(0.4%) | 190<br>(76.0%) | 81.4%<br>(69.6%-89.3%) | 99.5%<br>(97.1%-100%) |
| **Sampling strategy - Information available for N=1108** |  |  |  |  |  |  |  |
| **NP swab** | 1034<br>(93.3%) | 91<br>(8.8%) | 13<br>(1.3%) | 1<br>(0.1) | 929<br>(89.8%) | 87.5%<br>(79.8%-92.5%) | 99.9%<br>(99.4%-100%) |
| **OP swab** | 74<br>(6.7%) | 1<br>(1.4%) | 1<br>(1.4%) | 0 | 72<br>(97.3%) | 50.0%<br>(25.6%-97.4%) | 100%<br>(94.9%-100%) |
| **Symptom duration – Information available for N=687** |  |  |  |  |  |  |  |
| **0-7 days**<br>**Overall** | 610<br>(88.8%) | 69<br>(11.3%) | 7<br>(1.1%) | 1<br>(0.2%) | 533<br>(87.4%) | 90.8%<br>(82.2%-95.5%) | 99.6%<br>(98.9%-100%) |
| **8-14 days**<br>**Overall** | 70<br>(10.2%) | 8<br>(11.4%) | 5<br>(7.1%) | 0 | 57<br>(81.4%) | 61.5 %<br>(35.5%-82.3%) | 100%<br>(93.7%-100%) |
| **Symptomatic versus Asymptomatic – Information available for N=1100** |  |  |  |  |  |  |  |
| **Symptomatic** | 712<br>(64.7%) | 79<br>(11.1%) | 12<br>(1.7%) | 1<br>(0.1%) | 620<br>(87.1%) | 86.8%<br>(78.2%-92.3%) | 99.8%<br>(99.1%-100%) |
| **Asymptomatic** | 388<br>(35.3%) | 12<br>(3.1%) | 2<br>(0.5%) | 0 | 374<br>(96.4%) | 85.7%<br>(60.1%-96.0%) | 100%<br>(99.0%-100%) |
| **CT-Value PCR <30 and >=30 – Information available for N=105\*** |  |  |  |  |  |  |  |
| **CT value PCR**<br>**<30** | 93<br>(88%) | 87<br>(93.5%) | 6<br>(6.5%) | NA | NA | 93.5%<br>(86.6%-97.0%) | NA |
| **CT value PCR**<br>**>=30** | 12<br>(11.0%) | 4<br>(33.0%) | 8<br>(67.0%) | NA | NA | 33%<br>(13.8%-60.1%) | NA |
| **CT-Value PCR <25 and >=25 – Information available for N=105\*** |  |  |  |  |  |  |  |
| **CT value PCR**<br>**<25** | 72<br>(68.0%) | 69<br>(95.8%) | 3<br>(4.2%) | NA | NA | 95.8%<br>(88.5%-98.6%) | NA |
| **CT value PCR**<br>**>=25** | 33<br>(31.0%) | 22<br>(66.7%) | 11<br>(33.3%) | NA | NA | 66.7%<br>(49.6%-80.2%) | NA |

\*For one positive RT-PCR case CT-Value/Viral Load not provided due to a mutation in the E-Gene

**Figure 2:**
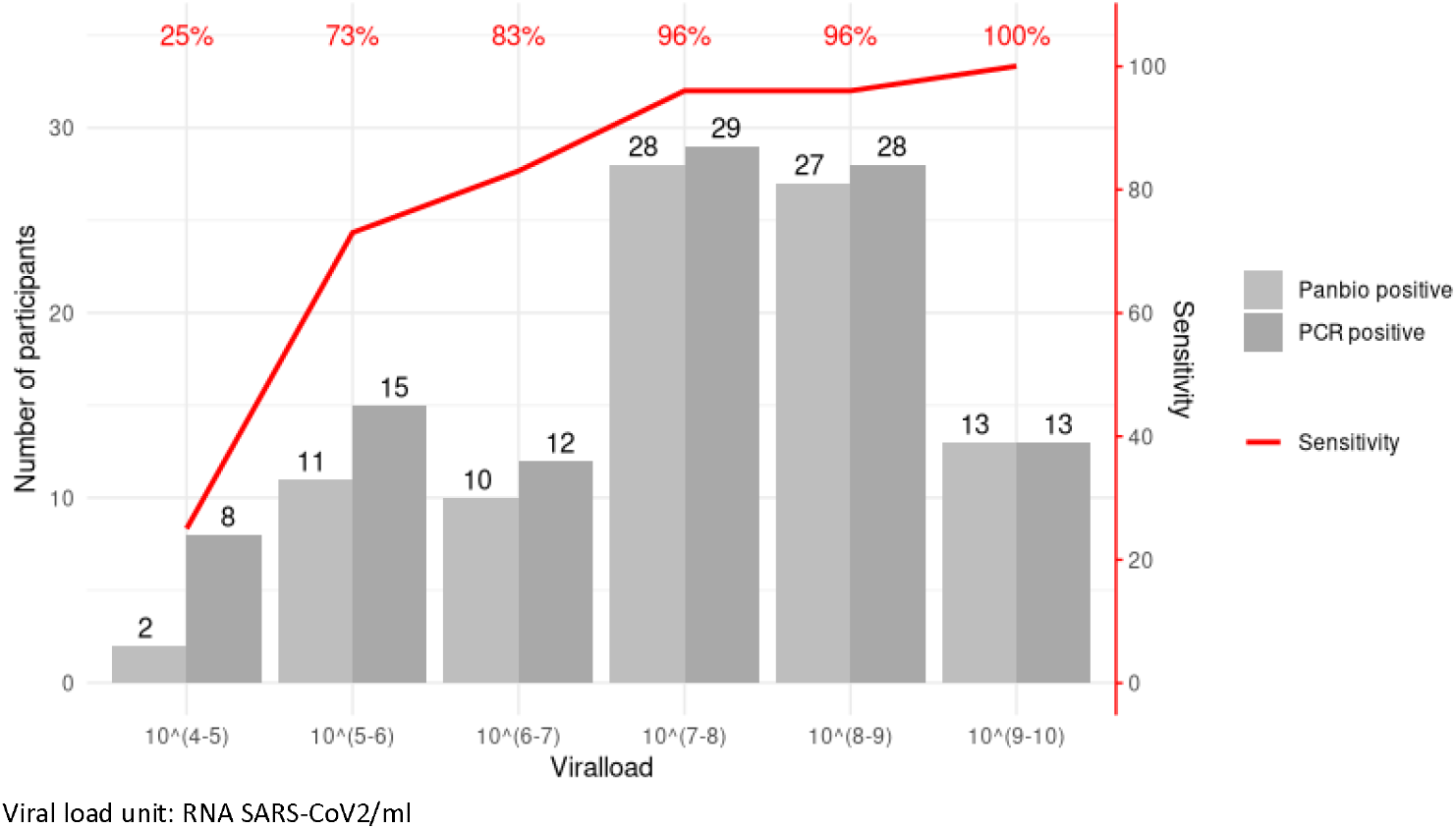
Sensitivity of PanBio Ag-RDT compared to viral load for all PCR positive cases (105 participants)

When assessing test performance by duration of symptoms, we found PanBio performed well in the first 7 days after symptom onset (sensitivity 90∙8% (95% CI: 82∙2%-95∙5%)), with declining sensitivity thereafter (>7 days of symptoms, sensitivity 61∙5 % (95% CI: 35∙5%-82∙3%)). This decrease in sensitivity with prolonged symptom duration is also shown in Figure 3, presenting the performance of PanBio with increasing days since symptom onset in relation to the calculated viral load for symptomatic and asymptomatic participants and the Ag-RDT performance.

**Figure 3:**
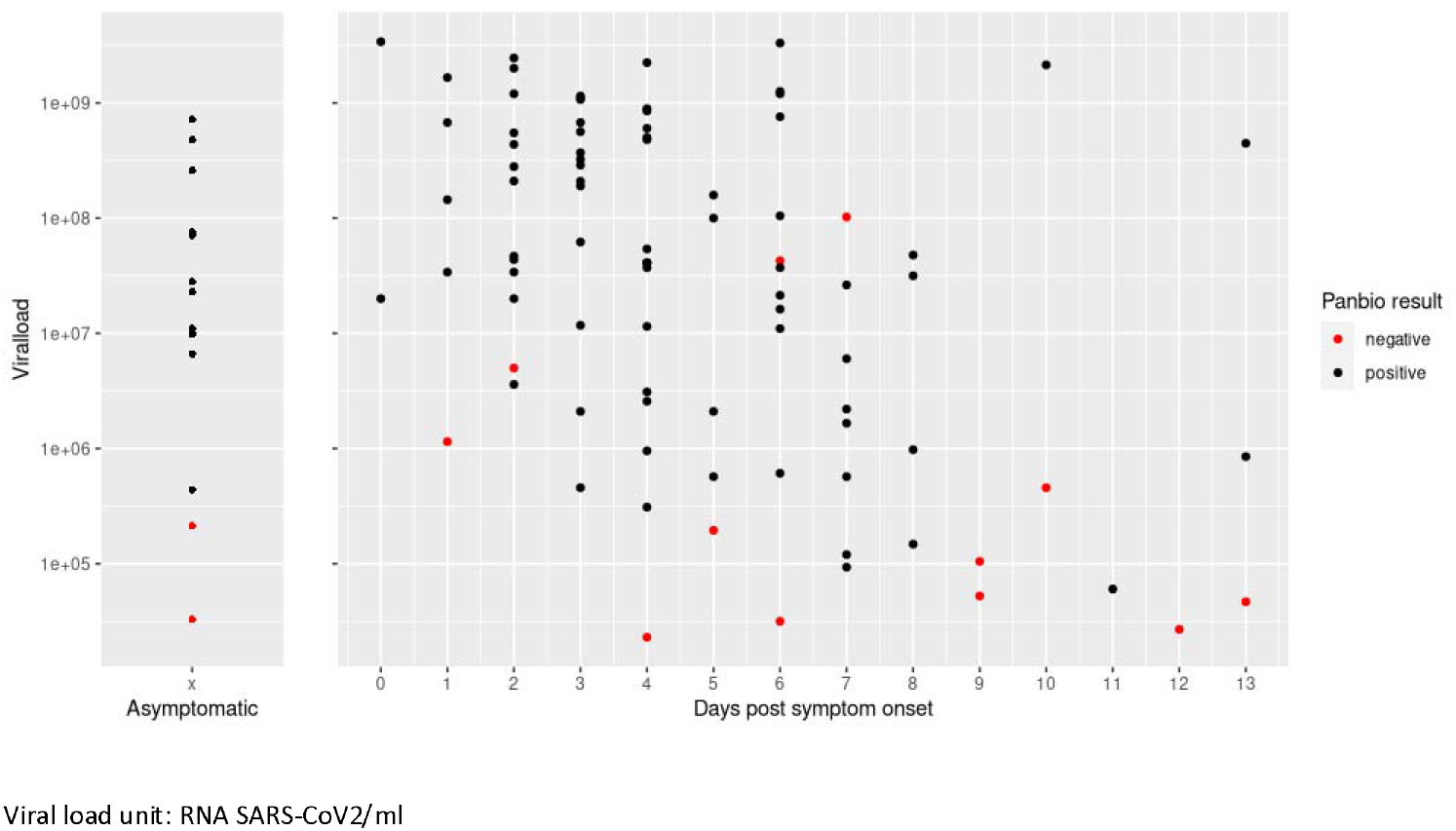
Viral load and Ag-RDT results for asymptomatic participants and by days post symptom onset for all PCR positive cases (105 participants)

Out of the total 106 positive RT-PCR cases, 14 participants were asymptomatic high-risk contacts. Within this small participant group the sensitivity of the Ag-RDT was 85∙7% (60∙1%-96∙0%), which compares to the sensitivity of symptomatic participants at 86∙8% (78∙2%-92∙3%); Table 2). Mean CT value in asymptomatic was 22∙1 (SD 4∙4) versus 23∙1 (SD 5∙0) in symptomatic participants.

The interrater reliability with kappa of 0.988 suggests that the tests results are clearly interpretable. PanBio scored 86 out of 100 points in the SUS showing a test which is easy to use. Problems were encountered when applying the exact amount of the required five drops to the test device, in addition to the handling of the buffer solution, meaning the squeezing to apply an appropriate amount onto the cassette formatted test device, which was considered tedious (Figure 4. System Usability Score and Ease-of-Use assessment results).

**Figure 4:**
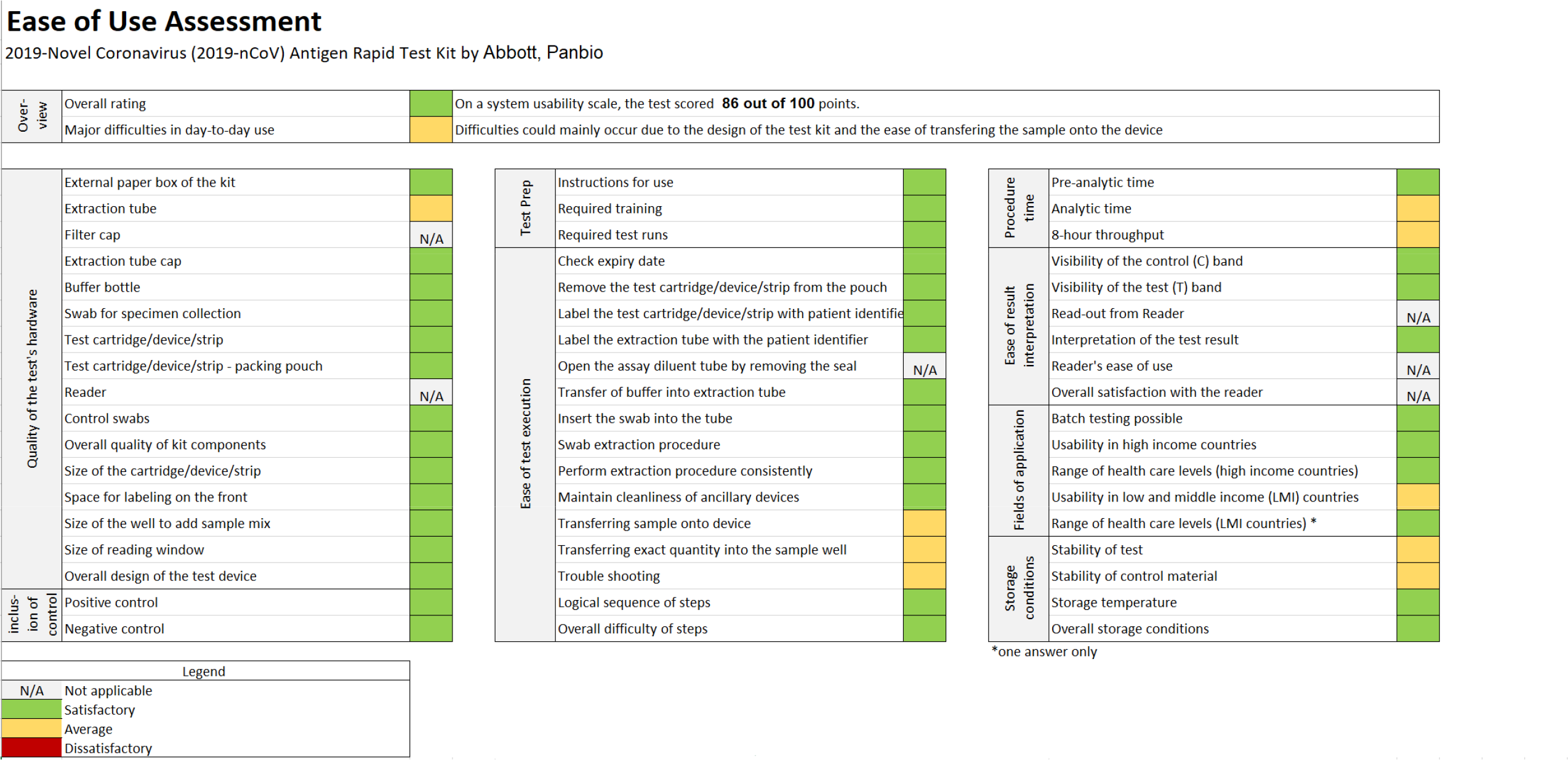
System Usability Score and Ease-of-Use assessment results. More explanation on the detailed EoU assessment is available in supplement material section (D).

## Discussion

This prospective multi-centre clinical diagnostic accuracy study shows that the PanBio Ag-RDT from Abbott has a high sensitivity of 86∙8% and an excellent specificity of 99∙9 % compared to the reference standard RT-PCR, and is easily performed in a point-of-care setting.

The differences observed in sensitivity between the two enrolment sites are probably explained by the different stages of the pandemic control. Berlin had a substantially higher prevalence with primarily symptomatic patients being tested and testing occurring later in the disease. Although the overall viral load on average was the same, the distribution of the viral load was not the same and more participants presented with low viral loads in Berlin, often late in the disease. There were 15 patients presenting with viral loads <6 log10 copies/mL versus only 8 in Heidelberg. The samples with low viral loads, were responsible for 8 out of the 11 false-negative results in Berlin, and 2 out of 3 in Heidelberg.

With a performance of 93∙5% for CT value <30 and 90∙8% within the first 7 days of symptoms, the test is likely to detect a large amount of transmission relevant SARS-CoV2 infections supporting recent published literature.^18,19^ Within the limitation of what can be concluded due to the small sample size of asymptomatic participants and the fact that these participants were asymptomatic high-risk contacts and presenting early in the disease, the performance of the test was as good as in symptomatic patients with a sensitivity respectively of 85∙7% versus 86∙8%. This suggests the Panbio to be an option for screening independent of symptoms and is in line with recent data suggesting that viral load in adults does not differ between asymptomatic and symptomatic infections.^20^ Further research with a larger asymptomatic cohort is needed to confirm our findings, however a truly representative sample of asymptomatic patients would only be possible in a large surveillance study.

Considering the test’s ease-of-use and the rapid turn-around time between 15 and 20 minutes, along with its high specificity, it could be considered for several use-cases: (1) screening of patients in advance of admission for elective procedures; (2) screening in advance of events at high-risk of transmission (e.g. aggregated settings where contact cannot be avoided); or (3) planned encounters with persons at high-risk for severe disease of *SARS-CoV-2* (e.g. visitor in nursing homes) in addition to (4) the use in symptomatic patients when PCR is not available or together with PCR, when a rapid decision is necessary.

Furthermore, given that supervised self-sampling from the anterior nose is a reliable alternative to professional nasopharyngeal sampling,^21,22^ scale-up of testing appears possible without requiring large numbers of trained health-care workers.

Overall, our study has several strengths. The population enrolled for testing was representative of the pandemic observed in adults in Germany with a broad spectrum of clinical presentations (from asymptomatic with high-risk contacts to severely ill). Due to the wide-spread testing available and the good test and trace capabilities, the population tested is expected to be a representative spectrum of disease. Also, the tests were performed at POC thus mimicking the real-world challenges of POC testing. And lastly, the comprehensive ease-of-use assessment with a standardized SUS-tool and a questionnaire, developed specifically for the study, highlighted important points for operationalization of the test.

However, the study also has several limitations. First, it was conducted only in one country, thus making it less representative of the pandemic at large. Second, the reference standard testing was performed on an NP swab in Heidelberg versus an NP/OP swab in Berlin. However, a recent systematic review does not suggest those sampling methods to yield different results.^23^ And lastly, we performed difference PCR methods as a reference standard. However, we ensured comparability by calibrating the methods and reporting on viral load.

In summary, the favourable ease-of-use results and the limited infrastructure required for the Ag-RDT testing procedure, its high specificity in addition to the high sensitivity of the test in persons with high viral load, can empower control of population transmission if implemented in well-designed testing programs.^24-26^ Policy makers should move from considering only test sensitivity to more holistic testing strategies, incorporating Ag-RDTs in addition to and in combination with RT-PCR to optimize the reach and depth of testing. The biggest limitation to such strategies on a global scale are likely to be manufacturing production shortages. However, with two tests (SD Biosensor/Roche Standard Q and Abbott PanBio) from large manufacturers already showing very high sensitivity and being WHO recommended, many other Ag-RDT are likely to follow in the pipeline. The hope is justified that the demand for Ag-RDTs will be met and also that prices of the tests will fall as has been observed for RDTs for other diseases as for example Malaria.^4,27,28^

## Data Availability

Anonymised data is available upon request from the corresponding author

https://apps.who.int/trialsearch/Trial2.aspx?TrialID=DRKS00021220

## References

1. World Health Organization. Director-General’s opening remarks at the media briefing on COVID-19. 16.03.2020 2020. https://www.who.int/dg/speeches/detail/who-director-general-s-opening-remarks-at-the-media-briefing-on-covid-1916-march-2020 (accessed 10.11.2020).

2. Wolrd Health Organization. Antigendetection in the diagnosis of SARS-CoV-2 infection using rapid immunoassays Interim Guidance https://www.who.int/publications/i/item/antigen-detection-in-the-diagnosis-of-sars-cov-2infection-using-rapid-immunoassays, 2020.

3. Foundation of Innovative New Diagnsotics. SARS-CoV-2 diagnostic pipeline. 2020 https://www.finddx.org/covid-19/pipeline/?avance=all&type=Rapid+diagnostic+tests&test_target=Antigen&status=all&section=show-all&action=default (accessed 10.11.2020).

4. Krüger LJ, Gaeddert M, Köppel L, et al. Evaluation of the accuracy, ease of use and limit of detection of novel, rapid, antigen-detecting point-of-care diagnostics for SARS-CoV-2. medRxiv 2020: 2020.10.01.20203836.

5. Organisation WH. WHO Emergency Use Listing for In vitro diagnostics (IVDs) Detecting SARS-CoV-2, 2020.

6. Organisation WH. WHO Emergency Use Assessment Coronavirus disease (COVID-19) IVDs PUBLIC REPORT, 2020.

7. Berger A, Ngo Nsoga M-T, Perez Rodriguez FJ, et al. Diagnostic accuracy of two commercial SARS-CoV-2 Antigen-detecting rapid tests at the point of care in community-based testing centers. medRxiv 2020: 2020.11.20.20235341.

8. Alemany A, Baro B, Ouchi D, et al. Analytical and Clinical Performance of the Panbio COVID-19 Antigen-Detecting Rapid Diagnostic Test. medRxiv 2020: 2020.10.30.20223198.

9. Gremmels H, Winkel BMF, Schuurman R, et al. Real-life validation of the Panbio COVID-19 Antigen Rapid Test (Abbott) in community-dwelling subjects with symptoms of potential SARS-CoV-2 infection. medRxiv 2020: 2020.10.16.20214189.

10. Denkinger CM, Brümmer L Diagnostics Global Health 2020 https://diagnosticsglobalhealth.org/.

11. Organisation WH. SARS-CoV-2 Antigen detecting rapid diagnostic test implementation projects. 10.11.2020 2020. https://www.who.int/news-room/articles-detail/sars-cov-2-antigen-detecting-rapid-diagnostic-test-implementation-projects (accessed 23.11.2020).

12. Abbott. PanBio Covid-19 Ag Rapid Test Device Instruction for Use 2020.

13. Corman VM, Landt O, Kaiser M, et al. Detection of 2019 novel coronavirus (2019-nCoV) by real-time RT-PCR. Euro Surveill 2020; 25(3).

14. Harris PA, Taylor R, Thielke R, Payne J, Gonzalez N, Conde JG. Research electronic data capture (REDCap)--a metadata-driven methodology and workflow process for providing translational research informatics support. J Biomed Inform 2009; 42(2): 377-81.

15. Harris PA TR, Thielke R, Payne J, Gonzalez N, Conde JG. A metadata-driven methodology and workflow process for providing translational research informatics support. J Biomed Inform 2009; 42(2): 377-81.

16. Bangor A KP, Miller JT. An Empirical Evaluation of the System Usability Scale. Intl Journal of Human–Computer Interaction 2008; 24: 574–94..

17. Altman DG, Bland JM. Diagnostic tests. 1: Sensitivity and specificity. BMJ 1994; 308(6943): 1552.

18. Jefferson T, Spencer, E., Brassey, J., Heneghan, C Viral cultures for COVID-19 infectivity assessment. Systematic review. medRix 2020.

19. Bulilete O, Lorente P, Leiva A, et al. Evaluation of the Panbio™ rapid antigen test for SARS-CoV-2 in primary health care centers and test sites. medRxiv 2020: 2020.11.13.20231316.

20. Kissler SM, Fauver JR, Mack C, et al. Viral dynamics of SARS-CoV-2 infection and the predictive value of repeat testing. medRxiv 2020: 2020.10.21.20217042.

21. Lindner AK, Nikolai O, Kausch F, et al. Head-to-head comparison of SARS-CoV-2 antigen-detecting rapid test with self-collected anterior nasal swab versus professional-collected nasopharyngeal swab. medRxiv 2020: 2020.10.26.20219600.

22. Abdulrahman A, Mustafa F, AlAwadhi AI, Alansari Q, AlAlawi B, AlQahtani M. Comparison of SARS-COV-2 nasal antigen test to nasopharyngeal RT-PCR in mildly symptomatic patients. medRxiv 2020: 2020.11.10.20228973.

23. Lee RA, Herigon JC, Benedetti A, Pollock NR, Denkinger CM. Performance of Saliva, Oropharyngeal Swabs, and Nasal Swabs for SARS-CoV-2 Molecular Detection: A Systematic Review and Meta-analysis. medRxiv 2020: 2020.11.12.20230748.

24. Larremore DB, Wilder B, Lester E, et al. Test sensitivity is secondary to frequency and turnaround time for COVID-19 surveillance. medRxiv 2020: 2020.06.22.20136309.

25. van Beek J, Igloi Z, Boelsums T, et al. From more testing to smart testing: data-guided SARS-CoV-2 testing choices. medRxiv 2020: 2020.10.13.20211524.

26. Paltiel AD, Zheng A, Walensky RP. Assessment of SARS-CoV-2 Screening Strategies to Permit the Safe Reopening of College Campuses in the United States. JAMA Netw Open 2020; 3(7): e2016818.

27. World Health Organisation. Advice on the use of point-of-care immunodiagnostic tests for COVID-19. 08.04.2020 2020 https://www.who.int/news-room/commentaries/detail/advice-on-the-use-of-point-of-care-immunodiagnostic-tests-for-covid-19 (accessed 26.08.2020).

28. Foundation of Innovative New Diagnostics. FIND SARS-COV-2 Diagnostics Pipeline 2020 https://www.finddx.org/covid-19/pipeline/ (accessed 30.09.2020).

