## Supplementary material for "Evaluation of the accuracy and ease-of-use of Abbott PanBio - A WHO emergency use listed, rapid, antigen-detecting point-of-care diagnostic test for *SARS-CoV-2*": Evaluation of the accuracy and ease of use of PanBio_Supplement for submission.pdf (from another version of this preprint: version 1, 2020-11-30)

**(A) Table 1: Study Team**

| | |
|---|---|
| Department of Public Health Rhein Neckar Region, Heidelberg, Germany | Dr. Kholoud Assaad, |
| | Dr. Andrea Fuhs |
| | Dr. Christoph Harter |
| | Christoph Schulze |
| | Gunter Schmitt |
| Division of Clinical Tropical Medicine, Heidelberg University Hospital, Germany | Anja Klemmer |
| | Lisa Köppel |
| | Maximilian Schirmer |
| | Lukas Brümmer |
| | Matthias Meinlschmidt |
| | Valerie Dürr |
| | Sofie Knoll |
| | Ann-Kathrin Backes |
| | Salome Steinke |
| | Alexander Penning |
| | Vanda Stankovic |
| | Henrik Ellinghaus |
| | Alina Schuckert |
| | Loai Abutaima |
| Institute of Tropical Medicine and International Health, Charité – Universitätsmedizin Berlin, Berlin, Germany | Chiara Rohardt |
| | Mia Wintel |
| | Franka Kausch |
| | Claudia Hülso |
| | Elisabeth Linzbach |
| Medical Directorate, Charité – Universitätsmedizin Berlin, Berlin, Germany | Heike Rössig |
| Institute of Tropical Medicine and International Health, Charité – Universitätsmedizin Berlin, Berlin, Germany | Maximilian Gertler |
| Charité Comprehensive Cancer Center, Charité – Universitätsmedizin Berlin, Berlin, Germany | Susen Burock |
| Department of Pediatric Surgery, Charité – Universitätsmedizin Berlin, Berlin, Germany | Katja von dem Busche |
| Berlin Institute for Clinical Teratology and Drug Risk Assessment in Pregnancy, Institute of Clinical Pharmacology and Toxicology, Charité – Universitätsmedizin Berlin, Berlin, Germany | Stephanie Padberg |

**(B) Section: Questionnaire for study participants**

**We invite you to participate in this survey. The survey serves to understand the diagnostic process and the disease and factors related to SARS-CoV-2 (novel coronavirus) infection.**

**Your answers will be kept strictly confidential and will not have a negative impact on your care. Participation in the study is voluntary and you have the option to skip questions that you do not want to answer.**

**The survey is expected to take 15-20 minutes. Thank you for your understanding and cooperation!**

| | |
|---|---|
| Postal code | *(Free text)* |
| Gender | ○ Male<br>○ Female<br>○ Diverse |
| How tall are you (in centimetres)? | *(Free text)* |
| How much do you weight (in kilograms)? | *(Free text)* |

**Symptoms that you attribute to the possible COVID-19**

| | |
|---|---|
| Did you have any symptoms of possible COVID-19 on the day of the test? | ○ No<br>○ Yes |
| Increased temperature / fever? | ○ No<br>○ Yes |
| Did you measure your fever? | ○ No<br>○ Yes |
| Highest temperature (in Celsius) | *(Free text)* |
| Cough | ○ No<br>○ Yes |
| Do you have a productive cough? | ○ No<br>○ Yes |
| Sore throat | ○ No<br>○ Yes |
| Shortness of breath | ○ No<br>○ Yes |
| Muscle pain / Body aches | ○ No<br>○ Yes |
| Fatigue | ○ No<br>○ Yes |

| | |
|---|---|
| Headache | ○ No<br>○ Yes |
| Runny nose | ○ No<br>○ Yes |
| Chest pain | ○ No<br>○ Yes |
| Diarrhea | ○ No<br>○ Yes |
| Nausea / vomiting | ○ No<br>○ Yes |
| Loss of taste or smell | ○ No<br>○ Yes |
| Other | ○ No<br>○ Yes |
| If yes, please specify | *(Free text)* |
| The earliest onset of symptoms attributed to possible COVID-19 | *(Day / Month / Year)* |
| How sick did you feel on the day of the test? | ○ Normal unrestricted activity as before the illness<br>○ Restriction with physical exertion, but able to walk; light physical work or work while sitting, e.g. light housework or office work, possible<br>○ Able to walk, self-sufficiency possible, but not able to work; can get up more than 50% of the waking time<br>○ Only limited self-sufficiency possible; 50% or more of the waking time tied to bed or chair<br>○ Completely in the need of care, no self-sufficiency possible; completely tied to bed or chair |
| Did you previously test negative? | ○ No<br>○ Yes |
| If yes, when | *(Day / Month / Year)* |
| If yes, where | ○ University clinic – inpatient<br>○ University clinic – outpatient<br>○ Drive-in<br>○ Other |
| Other, please specify | *(Free text)* |

| | |
|---|---|
| Do you know where you might have been infected with COVID-19? | ○ Household contact<br>○ Social contact<br>○ Work contact<br>○ Contact in university / school / kindergarten from you or a child in the family<br>○ Travel to risk area<br>○ Do not know<br>○ Other |
| Risk area | *(Free text)* |
| Other: please describe | *(Free text)* |

**Do you have any pre-existing conditions?**

| | |
|---|---|
| Which of the following lung disease(s) do you have? | ○ Asthma<br>○ Chronic Obstructive Pulmonary Disease – COPD<br>○ Breathing disorders during sleep<br>○ Obstructive Sleep Apnea – OSAS<br>○ Interstitial Lung Disease<br>○ Lung Cancer<br>○ Other<br>○ None |
| What other lung diseases do you have? | *(Free text)* |
| Cardiovascular diseases (e.g. hypertension, stroke, etc.) | ○ No<br>○ Yes |
| Chronic kidney disease | ○ No<br>○ Yes |
| Diabetes | ○ No<br>○ Yes |
| Autoimmune Disease (e.g. Rheumatoid Arthritis, MS) | ○ No<br>○ Yes |
| HIV | ○ No<br>○ Yes |
| Overweight | ○ No<br>○ Yes |
| Other, please specify | *(Free text)* |

**(C) Section: System Usability Scale**

### Useability evaluation - part I

"Evaluation of the performance of novel rapid diagnostics

for SARS-CoV-2 at point-of-care"

#### *System Usability Scale (SUS)*

© Digital Equipment Corporation, 1986 adapted format, version 1.0: 02/05/20

Name of the test: ________________________________________________

User identifier and study site: __________________________ Date: _______________

| | Strongly disagree | | | | Strongly agree |
|---|---|---|---|---|---|
| 1. I think that I would like to use this system frequently | 1 | 2 | 3 | 4 | 5 |
| 2. I found the system unnecessarily complex | 1 | 2 | 3 | 4 | 5 |
| 3. I thought the system was easy to use | 1 | 2 | 3 | 4 | 5 |
| 4. I think that I would need the support of a technical person to be able to use this system | 1 | 2 | 3 | 4 | 5 |
| 5. I found the various functions in this system were well integrated | 1 | 2 | 3 | 4 | 5 |
| 6. I thought there was too much inconsistency in this system | 1 | 2 | 3 | 4 | 5 |
| 7. I would imagine that most people would learn to use this system very quickly | 1 | 2 | 3 | 4 | 5 |
| 8. I found the system very cumbersome to use | 1 | 2 | 3 | 4 | 5 |
| 9. I felt very confident using the system | 1 | 2 | 3 | 4 | 5 |
| 10. I needed to learn a lot of things before I could get going with this system | 1 | 2 | 3 | 4 | 5 |

##### *Using SUS*

The SU scale is generally used after the respondent has had an opportunity to use the system being evaluated, but before any debriefing or discussion takes place. Respondents should be asked to record their immediate response to each item, rather than thinking about items for a lang time.

All items should be checked. If a respondent feels that they cannot respond to a particular item, they should mark the centre point of the scale.

#### (D) Section: Ease-of-Use Assessment Questionnaire

#### Usability evaluation - part II

"Evaluation of the performance of novel rapid diagnostics for SARS-CoV-2 at point-of-care"

Thank you for your time to answer this questionnaire (about 20 minutes).

*Your input is very valuable!*

***OVERALL QUESTIONS***

1. **User identifier**
2. **Date of filling the questionnaire**
3. **In which country do you currently work?**
4. **At which facility / study site do you currently work?**

*Mark only one oval.*

- ◯ Reilingen (Heidelberg)
- ◯ Berlin
- ◯ Liverpool
- ◯ Macae CTC
- ◯ Marica (Guapi)
- ◯ UFRJ

5. **Which test are you assessing?**

*Mark only one oval.*

- ◯ Coris Bioconcept Respi Strip
- ◯ Bioeasy FIA
- ◯ Bioeasy Colloidal Gold
- ◯ SD Biosensor Standard F (Flourescence)
- ◯ SD Biosensor Standard Q
- ◯ Rapigen Biocredit Colloidal Gold
- ◯ Bionote, NowCheck
- ◯ Abbott Panbio

**6. About how many times did you perform this test approximately?**
*Mark only one oval.*

- ⬭ Only observed use
- ⬭ < 10
- ⬭ 10 – 100
- ⬭ > 100

**7. About how many times did you observe the use of this test (not performed yourself)?**
*Mark only one oval.*

- ⬭ < 10
- ⬭ 10 – 50
- ⬭ 50 – 100
- ⬭ > 100

**8. What is your profession?**

**9. How many years of laboratory experience do you have?**

**10. How many years of working experience in limited resource settings do you have?**

**11. How much experience do you have with interpreting the results of lateral flow tests or rapid diagnostics (e.g. for HIV, malaria, pregnancy)?** Please note that we refer here to your experience with INTERPRETING the test results. If you do not conduct the test yourself, but do inform patients about the test results, we also consider that as experience with INTERPRETING the test results.
*Mark only one oval.*

- ⬭ None
- ⬭ < 1 year
- ⬭ 1 – 3 years
- ⬭ > 3 years

#### *TEST SPECIFIC QUESTIONS*

##### *TRAINING*

**12. How satisfied were you with the following components of the test training?**
*Mark only one oval per row.*

| | Very satisfied | Satisfied | Neither | Dissatisfied | Very dissatisfied |
|---|---|---|---|---|---|
| Instructions for Use | ⬭ | ⬭ | ⬭ | ⬭ | ⬭ |
| Standard Operating Procedures | ⬭ | ⬭ | ⬭ | ⬭ | ⬭ |
| Face to face demonstration | ⬭ | ⬭ | ⬭ | ⬭ | ⬭ |

**13. What additional materials (if any) do you think should be provided as part of the training?**

None

Other: ______________________________________________

**14. How long should be the training of this test?**

*Mark only one oval.*

Self-explanatory, no need for training

1 - 2 hours

2 - 4 hours

Half a day

Full day

**15. Do you consider proficiency testing necessary?**

Proficiency testing as in assessing the user's performance or ability to run the test following the training.

*Mark only one oval.*

Yes

No

**16. Please comment here on the need for proficiency testing**

---

**17. After how many of these tests do you feel you could perform the test on your own (having access to the training material)?**

*Mark only one oval.*

1 – 2 tests

3 – 5 tests

6 – 10 tests

> 10 tests

*ASSESSMENT OF TEST*

**18. How satisfied are you with the quality of each of the components in the kit (in terms of ease of use and fit for purpose)?**

*Mark only one oval per row*

| | Very satisfied | Satisfied | Neither | Dissatisfied | Very dissatisfied |
|---|---|---|---|---|---|
| External paper box of kit | | | | | |
| Extraction tube | | | | | |
| Filter cap (if applicable) | | | | | |

| | | | | | |
|---|---|---|---|---|---|
| Extraction tube cap (if applicable) | ⬭ | ⬭ | ⬭ | ⬭ | ⬭ |
| Buffer bottle (if applicable) | ⬭ | ⬭ | ⬭ | ⬭ | ⬭ |
| Swab for specimen collection | ⬭ | ⬭ | ⬭ | ⬭ | ⬭ |
| Test cartridge / device | ⬭ | ⬭ | ⬭ | ⬭ | ⬭ |
| Test cartridge packing / pouch | ⬭ | ⬭ | ⬭ | ⬭ | ⬭ |
| Reader (if applicable) | ⬭ | ⬭ | ⬭ | ⬭ | ⬭ |
| Positive/negative control swabs (if applicable) | ⬭ | ⬭ | ⬭ | ⬭ | ⬭ |

**19. How useful do you find the inclusion of a positive control? (if applicable)**

| | 1 | 2 | 3 | 4 | 5 | |
|---|---|---|---|---|---|---|
| Very useful | ⬭ | ⬭ | ⬭ | ⬭ | ⬭ | Not useful |

**Please comment on the added value of a positive control to the test kit (if applicable)?**

______________________________________________________________________________

**20. How useful do you find the inclusion of a negative control? (if applicable)**

| | 1 | 2 | 3 | 4 | 5 | |
|---|---|---|---|---|---|---|
| Very useful | ⬭ | ⬭ | ⬭ | ⬭ | ⬭ | Not useful |

**Please comment on the added value of a negative control to the test kit (if applicable)?**

______________________________________________________________________________

**21. Which kit component(s) should be improved in your opinion (if any)?**
*Please specify why and how*

**22. Overall, how satisfied are you with the kit components?**
*Mark only one oval.*

| | 1 | 2 | 3 | 4 | 5 | |
|---|---|---|---|---|---|---|
| Very satisfied | ⬭ | ⬭ | ⬭ | ⬭ | ⬭ | Very dissatisfied |

**23. How satisfied are you with the overall design of the device in terms of the following features:**
*Mark only one oval per row.*

| | Very satisfied | Satisfied | Neither | Dissatisfied | Very dissatisfied |
|---|---|---|---|---|---|
| Size of cartridge | ⬭ | ⬭ | ⬭ | ⬭ | ⬭ |
| Space for labeling on the front | ⬭ | ⬭ | ⬭ | ⬭ | ⬭ |

| |
|---|
| Size of the well to add sample mix |
| Size of reading window |
| Logical sequence of steps |

**24. Overall, how satisfied are you with the design of the device?**

*Mark only one oval.*

| | 1 | 2 | 3 | 4 | 5 | |
|---|---|---|---|---|---|---|
| Very satisfied | | | | | | Very dissatisfied |

**25. Please assess the Test's storage conditions**

*Mark only one oval.*

| | > 12 months | 12 to 6 months | 5 to 3 months | < 3 months |
|---|---|---|---|---|
| Stability of test | | | | |
| Stability of control material (if appl.) | | | | |

| | 2 – 30° | 15 – 35° | 15 – 30° | 20 – 25° |
|---|---|---|---|---|
| Storage temperature | | | | |

**26. Overall, how satisfied are you with the test's storage conditions?**

*Mark only one oval.*

| | 1 | 2 | 3 | 4 | 5 | |
|---|---|---|---|---|---|---|
| Very satisfied | | | | | | Very dissatisfied |

**27. Which part(s) of the device could be improved in your opinion (if any)?**

*Please specify why and how.*

**28. Please determine the difficulty of the following steps :**

Please consider your day-to-day/routine workload (or that of the people in the lab/area where this test could be implemented) to answer this question. Please leave any steps blank if not applicable.

*Mark only one oval per row.*

| | Very easy | Easy | Neither | Difficult | Very difficult |
|---|---|---|---|---|---|

| | | | | | |
|---|---|---|---|---|---|
| a) Check expiry date | ◯ | ◯ | ◯ | ◯ | ◯ |
| b) Remove the test cartridge from the pouch | ◯ | ◯ | ◯ | ◯ | ◯ |
| c) Label the test cartridge with patient identifier | ◯ | ◯ | ◯ | ◯ | ◯ |
| d) Label the assay diluent tube with patient identifier | ◯ | ◯ | ◯ | ◯ | ◯ |
| e) Open the assay diluent tube by removing the seal (if appl.) | ◯ | ◯ | ◯ | ◯ | ◯ |
| f) Transfer of buffer into diluent tube (if applicable) | ◯ | ◯ | ◯ | ◯ | ◯ |
| g) Insert the swab into the tube | ◯ | ◯ | ◯ | ◯ | ◯ |
| h) Ease of swab extraction procedure | ◯ | ◯ | ◯ | ◯ | ◯ |
| i) Ability to perform swab extraction procedure consistently | ◯ | ◯ | ◯ | ◯ | ◯ |
| j) Ability to maintain cleanliness of ancillary devices (e.g. pipette) in order to avoid cross contamination | ◯ | ◯ | ◯ | ◯ | ◯ |
| k) Ease of transferring sample onto device | ◯ | ◯ | ◯ | ◯ | ◯ |
| l) Ease of transferring exact quantity into the sample well | ◯ | ◯ | ◯ | ◯ | ◯ |
| m) Trouble shooting | ◯ | ◯ | ◯ | ◯ | ◯ |

**29. How satisfied are you with the logical sequence of steps?**

*Mark only one oval.*

| | 1 | 2 | 3 | 4 | 5 | |
|---|---|---|---|---|---|---|
| Very satisfied | ◯ | ◯ | ◯ | ◯ | ◯ | Very dissatisfied |

**30. Overall, how difficult did you find the steps?**

*Mark only one oval.*

| | 1 | 2 | 3 | 4 | 5 | |
|---|---|---|---|---|---|---|
| Very easy | ◯ | ◯ | ◯ | ◯ | ◯ | Very difficult |

**31. Overall, how satisfied are you with the time relevant components?**

*Mark only one oval.*

| 1 | 2 | 3 | 4 | 5 |
|---|---|---|---|---|

| | | | | | | |
|---|---|---|---|---|---|---|
| Very satisfied | ⬭ | ⬭ | ⬭ | ⬭ | ⬭ | Very dissatisfied |

**32. Please assess the time relevant components of the test.**

*Mark only one oval per row.*

| | ≤ 2 min | 3 to 5 min | 6 to 10 min | > 10 min |
|---|---|---|---|---|
| Pre analytic time | ⬭ | ⬭ | ⬭ | ⬭ |
| Analytic time | ⬭ | ⬭ | ⬭ | ⬭ |

**33. In your opinion, about how many patients could be tested with this test in an 8-hour day?**

*Mark only one oval per row.*

- ⬭ < 10
- ⬭ 10 – 50
- ⬭ 50 – 100
- ⬭ > 100

**34. Please comment here if you see any potential issues or room for improvement.**

*<u>READ-OUT of TEST</u>*

**35. How did you find the results read-out in the following areas:**

*Mark only one oval per row.*

| | Very easy | Easy | Neither | Difficult | Very difficult |
|---|---|---|---|---|---|
| a) Visibility of the control (C) and in contrast with the background (if applicable)? | ⬭ | ⬭ | ⬭ | ⬭ | ⬭ |
| b) Visibility of the test (T) band in contrast with the background (if applicable)? | ⬭ | ⬭ | ⬭ | ⬭ | ⬭ |
| c) Read-out from Reader (if applicable) | ⬭ | ⬭ | ⬭ | ⬭ | ⬭ |
| d) Interpretation of the test result | ⬭ | ⬭ | ⬭ | ⬭ | ⬭ |

**36. Do you foresee any issues with reading these results considering the lighting conditions in the settings you currently work or have experience with?**

*Mark only one oval.*

- ⬭ Yes (please explain below)
- ⬭ No

**If yes, please explain here**

**37. For visual readout: Was there any color on the background of the test result (T) or control band (C) that made the interpretation of the bands difficult?**

*Mark only one oval.*

- Not applicable
- Yes (please explain below)
- No

**If yes, which background color was present?**

______________________________________________________________________________

**38. How satisfied are you with the reader in terms of ease of use and fit for purpose? If not applicable, please leave blank**

| | 1 | 2 | 3 | 4 | 5 | |
|---|---|---|---|---|---|---|
| Very satisfied | ⬭ | ⬭ | ⬭ | ⬭ | ⬭ | Very dissatisfied |

**39. Overall, how satisfied are you with the reader? If not applicable, please leave blank**

| | 1 | 2 | 3 | 4 | 5 | |
|---|---|---|---|---|---|---|
| Very satisfied | ⬭ | ⬭ | ⬭ | ⬭ | ⬭ | Very dissatisfied |

**40. Which component(s) of the reader (if applicable) should be improved in your opinion (if any)?**
*Please specify why and how*

---

*<u>OVERALL ASSESMMENT</u>*

**41. Overall, how did you find the use of this rapid COVID-19 diagnostic tool:**
*Mark only one oval.*

| | 1 | 2 | 3 | 4 | 5 | |
|---|---|---|---|---|---|---|
| Very easy | ⬭ | ⬭ | ⬭ | ⬭ | ⬭ | Very difficult |

**42. Please comment here on the use:**

---

**43. Which option(s) do you consider feasible in your setting?**

- Sequential testing (run tests one by one) ONLY
- Sequential testing AND batch testing (run multiple tests at the time)

***44.*** **Which aspect(s) of this test could cause difficulties in its day-to-day use?** *Tick all that apply.*

- Hands-on time
- Tatal assay time to result
- Batch processing
- Throughput
- Test results interpretation
- Overall number of steps
- Time sensitive steps
- Cartridge design
- Quality of material
- Training requirements
- Storage conditions and stability
- Waste management requirements
- I don’t know
- None, I see no barriers for implementation

**45. Please give a short explanation for each of the aspects you selected above e.g. what could be the challenges in the day-to-day use:**

---

*<u>SETTINGS UF USE</u>*

**46. Do you see this test being used in its current form in your setting in your country?**
*Mark only one oval.*

- Yes (please explain below)
- No (please explain below)
- I don’t know

Please elaborate:

---

**47. If yes, at which health care level(s) do you see this test being implemented in your country**
*Tick all that apply.*

- Family doctor / General physician
- Peripheral hospital / lab

- Reference hospital / lab
- At a testing site operated by traines staff without specific laboratory expertise

**48. If you don't see this test being used in its current form, which aspects should be changed to make it suitable for use in your setting in your country:**

**49. Do you see this test being used in its current form in your setting in LOW- and MIDDLE-INCOME COUNTRIES?**

- Yes (please explain below)
- No (please explain below)
- I don’t know

Please elaborate:

**50. At which health care level(s) do you see this test being implemented in low and middle income countries?**

- Family doctor / General physician
- Primary health care
- Health centre / microscopy lab
- District hospital / lab
- Reference hospital / lab
- Other:
- I cannot answer this questions as I have no work experience in those countris

**51. If you don't see this test being used in its current form, which aspects should be changed to make it suitable for use in your setting in low and middle income countries?**

**52. Anything else you would like to add?**

#### THANK YOU VERY MUCH!

#### (E) Figure 1: Interpretation matrix for Ease of Use Assessment

| # | Question | green | green | green | yellow | yellow | amber | amber |
|---|---|---|---|---|---|---|---|---|
| 1 | User Identifier (first name, surname) | - | - | - | - | - | - | - |
| 2 | Date of filling the questionnaire | - | - | - | - | - | - | - |
| 3 | In which country do you currently work? | - | - | - | - | - | - | - |
| 4 | At which facility / study site do you currently work? | - | - | - | - | - | - | - |
| 5 | Which test are you assessing? | - | - | - | - | - | - | - |
| 6 | About how many times did you perform this test approximately? | - | - | - | - | - | - | - |
| 7 | About how many times did you observe the use of this test (not performed yourself)? | - | - | - | - | - | - | - |
| 8 | What is your profession? | - | - | - | - | - | - | - |
| 9 | How many years of laboratory experience do you have? | - | - | - | - | - | - | - |
| 10 | How many years of working experience in limited resource settings do you have? | - | - | - | - | - | - | - |
| 11 | How much experience do you have with interpreting the results of lateral flow tests or rapid diagnostics (e.g. for HIV, malaria, pregnancy)? | - | - | - | - | - | - | - |
| 12 | How satisfied were you with the following components of the test training? [Instructions for use] | very satisfied | satisfied | - | neither | - | dissatisfied | very dissatisfied |
| 12 | How satisfied were you with the following components of the test training? [Standard operating procedures] | - | - | - | - | - | - | - |
| 12 | How satisfied were you with the following components of the test training? [Face - to - face demonstration] | - | - | - | - | - | - | - |
| 13 | What additional materials (if any) do you think should be provided as part of the training? | None | - | - | 1-2 | - | >2 | - |
| 14 | How long should the training of this test be? | Self - explanatory, no | 1 - 2 hours of training | 2 - 4 hours of training | Half a day | - | Full Day | - |
| 15 | Do you consider proficiency testing necessary? | No | - | - | - | - | Yes | - |
| 16 | Please comment here on the need for proficiency testing: | - | - | - | - | - | - | - |
| 17 | After how many of these tests do you feel you could perform the test on your own? | 1 - 2 tests | - | - | 3 - 5 tests | - | 6 - 10 tests | 10 tests |
| 18 | How satisfied are you with the quality of each of the components in the kit (in terms of ease of use and fit for purpose)? [External paper box of the kit] | very satisfied | satisfied | - | neither | - | dissatisfied | very dissatisfied |
| 18 | How satisfied are you with the quality of each of the components in the kit (in terms of ease of use and fit for purpose)? [Assay diluent tube] | very satisfied | satisfied | - | neither | - | dissatisfied | very dissatisfied |
| 18 | How satisfied are you with the quality of each of the components in the kit (in terms of ease of use and fit for purpose)? [Filter cap (if necessary)] | very satisfied | satisfied | - | neither | - | dissatisfied | very dissatisfied |
| 18 | How satisfied are you with the quality of each of the components in the kit (in terms of ease of use and fit for purpose)? [Swab for specimen collection] | very satisfied | satisfied | - | neither | - | dissatisfied | very dissatisfied |
| 18 | How satisfied are you with the quality of each of the components in the kit (in terms of ease of use and fit for purpose)? [Test cartridge - device] | very satisfied | satisfied | - | neither | - | dissatisfied | very dissatisfied |
| 18 | How satisfied are you with the quality of each of the components in the kit (in terms of ease of use and fit for purpose)? [Test cartridge - packing pouch] | very satisfied | satisfied | - | neither | - | dissatisfied | very dissatisfied |
| 18 | How satisfied are you with the quality of each of the components in the kit (in terms of ease of use and fit for purpose)? [Reader (if necessary)] | very satisfied | satisfied | - | neither | - | dissatisfied | very dissatisfied |
| 19 | Which kit component(s) should be improved in your opinion (if any)? | - | - | - | - | - | - | - |
| 20 | Overall, how satisfied are you with the kit components? | 1 | 2 | - | 3 | - | 4 | 5 |
| 21 | How satisfied are you with the design of the following features? [Size of the cartridge] | very satisfied | satisfied | - | neither | - | dissatisfied | very dissatisfied |
| 21 | How satisfied are you with the design of the following features? [Space for labeling on the front (e.g. patient ID)] | very satisfied | satisfied | - | neither | - | dissatisfied | very dissatisfied |
| 21 | How satisfied are you with the design of the following features? [Size of the well to add sample mix] | very satisfied | satisfied | - | neither | - | dissatisfied | very dissatisfied |
| 21 | How satisfied are you with the design of the following features? [Size of reading window] | very satisfied | satisfied | - | neither | - | dissatisfied | very dissatisfied |
| 21 | How satisfied are you with the design of the following features? [Logical sequence of steps] | - | - | - | - | - | - | - |
| 22 | Overall, how satisfied are you with the design of the device? | 1 | 2 | - | 3 | - | 4 | 5 |
| 23 | Which part(s) of the device could be improved in your opinion (if any)? | - | - | - | - | - | - | - |
| 24 | Please assess the test's storage conditions. [Stability of test] | 12 months | - | - | 12 to 6 months | 5 to 3 months | 3 months | - |
| 24 | Please assess the test's storage conditions. [Stability of control material (if applicable)] | 12 months | - | - | 12 to 6 months | 5 to 3 months | 3 months | - |
| 24 | Please assess the test's storage conditions. [Storage temperature] | 2 - 40° | - | - | 15- 35° | 15 - 30° | 20 - 25° | - |
| 25 | Overall, how satisfied are you with the test's storage conditions? | 1 | 2 | - | 3 | - | 4 | 5 |
| 26 | Please determine the difficulty of the following steps: [a) Check expiry date] | Very easy | Easy | - | Neither | - | Difficult | Very difficult |
| 26 | Please determine the difficulty of the following steps: [b) Remove the test cartridge from the pouch] | Very easy | Easy | - | Neither | - | Difficult | Very difficult |
| 26 | Please determine the difficulty of the following steps: [c) Label the test cartridge with the patient identifier] | Very easy | Easy | - | Neither | - | Difficult | Very difficult |
| 26 | Please determine the difficulty of the following steps: [d) Label the assay diluent tube with the patient identifier] | Very easy | Easy | - | Neither | - | Difficult | Very difficult |
| 26 | Please determine the difficulty of the following steps: [e) Open the assay diluent tube by removing the seal (if appl.)] | Very easy | Easy | - | Neither | - | Difficult | Very difficult |
| 26 | Please determine the difficulty of the following steps: [f) Transfer of buffer into diluent tube (if appl.)] | Very easy | Easy | - | Neither | - | Difficult | Very difficult |
| 26 | Please determine the difficulty of the following steps: [g) Insert the swab into the tube] | Very easy | Easy | - | Neither | - | Difficult | Very difficult |
| 26 | Please determine the difficulty of the following steps: [h) Ease of swab extraction procedure] | Very easy | Easy | - | Neither | - | Difficult | Very difficult |
| 26 | Please determine the difficulty of the following steps: [i) Ability to perform extraction procedure consistently] | Very easy | Easy | - | Neither | - | Difficult | Very difficult |
| 26 | Please determine the difficulty of the following steps: [j) Ability to maintain cleanliness of ancillary devices (e.g. pipette) in order to avoid cross-contamination] | Very easy | Easy | - | Neither | - | Difficult | Very difficult |
| 26 | Please determine the difficulty of the following steps: [k) Ease of transferring sample onto device] | Very easy | Easy | - | Neither | - | Difficult | Very difficult |
| 26 | Please determine the difficulty of the following steps: [l) Ease of transferring exact quantity into the sample well] | Very easy | Easy | - | Neither | - | Difficult | Very difficult |
| 26 | Please determine the difficulty of the following steps: [m) Trouble shooting] | Very easy | Easy | - | Neither | - | Difficult | Very difficult |
| 27 | How satisfied are you with the logical sequence of steps? | 1 | 2 | - | 3 | - | 4 | |
| 28 | Overall, how difficult did you find the steps? | 1 | 2 | - | 3 | - | 4 | 4 |
| 29 | Please assess the time relevant components of the test. [Pre-analytic time] | 2 min | - | - | 3 to 5 min | 6 to 10 min | 10 min | - |
| 29 | Please assess the time relevant components of the test. [Analytic time] | 2 min | - | - | 3 to 5 min | 6 to 10 min | 10 min | - |
| 30 | In your opinion, about how many patients could be tested with this test in an 8-hour day? | 100 | - | - | 50 - 100 | - | 10 - 50 | 10 |
| 31 | Please comment here if you see any potential issues or room for improvement. | - | - | - | - | - | - | - |
| 32 | How did you find the results read-out in the following areas: [a) Visibility of the control (C) band in contrast with the background (if appl.)] | Very easy | Easy | - | Neither | - | Difficult | Very difficult |
| 32 | How did you find the results read-out in the following areas: [b) Visibilty of the test (T) band in contrast with the background (if appl.)] | Very easy | Easy | - | Neither | - | Difficult | Very difficult |
| 32 | How did you find the results read-out in the following areas: [c) Read-out from Reader (if appl.)] | Very easy | Easy | - | Neither | - | Difficult | Very difficult |
| 32 | How did you find the results read-out in the following areas: [d) Interpretation of the test result] | Very easy | Easy | - | Neither | - | Difficult | Very difficult |
| 33 | Do you foresee any issues with reading these results considering the lighting conditions in the settings you currently work or have experience with? | - | - | - | - | - | - | - |
| 33 | Please elaborate | - | - | - | - | - | - | - |
| 34 | For visual readout: Was there any color on the background of the test result (T) or control band (C) that made the interpretation of the bands difficult? | - | - | - | - | - | - | - |
| 34 | If yes, which background color was present | - | - | - | - | - | - | - |
| 35 | How satisfied are you with the reader (if applicable) in terms of ease of use and fit for purpose? | 1 | 2 | - | 3 | - | 4 | 5 |
| 36 | Overall, how satisfied are you with the reader (if applicable)? | 1 | 2 | - | 3 | - | 4 | 5 |
| 37 | Which component(s) of the reader (if applicable) should be improved in your opinion (if any)? | - | - | - | - | - | - | - |
| 38 | Overall, how did you find the use of this rapid COVID-19 diagnostic tool? | 1 | 2 | - | 3 | - | 4 | 5 |
| 39 | Please comment here on the use: | - | - | - | - | - | - | - |
| 40 | Which option(s) do you consider feasible in your setting? | Batch testing (run mul | - | - | - | - | Sequential testing (rur | - |
| 41 | Which aspect(s) of this test could cause difficulties in its day-to-day use? | 0 | - | - | 1 | 2 | 3 | 4 |
| 42 | Please give a short explanation for each of the aspects you selected above e.g. what could be the challenges in the day-to-day use: | - | - | - | - | - | - | - |
| 43 | Do you see this test being used in its current form in your setting in your country? | Yes (please explain below) | | - | - | - | No (please explain bel | - |
| 43 | Please elaborate | - | - | - | - | - | - | - |
| 44 | If yes, at which health care level(s) do you see this test being implemented in your country? | At a testing site opera | Family doctor / Gener | - | Peripheral hospital / la | - | Reference hospital / la | - |
| 45 | If you don't see this test being used in its current form, which aspects should be changed to make it suitable for use in your setting in your country: | none | - | - | 1-2 | - | >2 | - |
| 46 | Do you see this test being used in its current form in your setting in LOW and MIDDLE INCOME COUNTRIES? | Yes (please explain be | - | - | - | - | No (please explain bel | - |
| 46 | Please elaborate | - | - | - | - | - | - | - |
| 47 | At which health care level(s) do you see this test being implemented in low and middle income countries? | Primary health care | Famildy doctor / Gene | Health centre / micros | District hospital / labo | - | Reference hospital / la | - |
| 48 | If you don't see this test being used in its current form, which aspects should be changed to make it suitable for use in your setting in low and middle income countries? | none | - | - | 1 | 2 | 3 | 4 |
| 49 | Anything else you would like to add? | - | - | - | - | - | - | - |

#### (F) Table 2: Detailed list of viral load and symptoms for all PCR positives

| Viral load* | Result Ag-RDT | Increased temperature/ Fever | Cough | Do you have a productive cough? | Sore throat | Shortness of breath | Muscle pain/ Body aches | Fatigue | Headache | Runny nose | Chest pain | Diarrhea | Nausea/ vomiting | Loss of taste or smell | Other |
|---|---|---|---|---|---|---|---|---|---|---|---|---|---|---|---|
| 7.53 | positive | No | Yes | Yes | Yes | No | Yes | Yes | Yes | No | Yes | No | No | No | No |
| 8.86 | positive | | | | | | | | | | | | | | |
| 9.53 | positive | Yes | No | No | Yes | No | Yes | Yes | Yes | No | No | No | No | No | No |
| 8.45 | positive | Yes | Yes | Yes | No | No | Yes | Yes | Yes | No | Yes | Yes | Yes | Yes | No |
| 8.41 | positive | | | | | | | | | | | | | | |
| 7.04 | positive | | | | | | | | | | | | | | |
| 6.32 | positive | No | No | No | No | No | No | No | No | Yes | No | No | No | No | No |
| 7.61 | positive | No | No | No | No | No | No | No | No | Yes | No | No | No | No | No |
| 8.68 | positive | No | Yes | No | No | No | Yes | No | No | No | Yes | Yes | No | No | No |
| 4.36 | negative | No | No | No | No | No | No | No | No | No | No | No | No | Yes | No |
| 8.32 | positive | Yes | Yes | Yes | No | No | Yes | Yes | Yes | No | No | No | No | No | No |
| 8.00 | positive | Yes | Yes | No | Yes | No | Yes | Yes | Yes | No | No | No | No | No | No |
| 9.04 | positive | Yes | Yes | No | No | No | Yes | Yes | Yes | Yes | No | No | No | No | No |
| 7.61 | positive | No | Yes | Yes | No | No | Yes | Yes | Yes | No | No | No | No | No | No |
| 8.74 | positive | No | | | Yes | | Yes | Yes | Yes | | | | | | |
| 7.61 | positive | Yes | Yes | No | No | Yes | Yes | Yes | Yes | No | No | Yes | No | No | No |
| 7.85 | positive | | | | | | | | | | | | | | |
| 8.68 | positive | | | | | | | | | | | | | | |
| 7.00 | positive | | | | | | | | | | | | | | |
| 7.88 | positive | | | | | | | | | | | | | | |
| 7.45 | positive | | | | | | | | | | | | | | |
| 9.08 | positive | No | Yes | No | No | No | No | No | No | Yes | No | No | No | No | No |
| 7.04 | positive | No | Yes | Yes | No | No | Yes | Yes | No | No | No | Yes | No | No | No |
| 5.76 | positive | No | Yes | Yes | No | No | Yes | Yes | Yes | No | No | No | Yes | Yes | |
| 6.34 | positive | No | Yes | No | No | No | No | No | No | Yes | No | No | No | No | No |
| 9.30 | positive | No | Yes | No | No | No | Yes | Yes | Yes | Yes | Yes | No | No | Yes | No |
| 6.56 | positive | No | Yes | Yes | Yes | No | Yes | No | No | Yes | No | No | No | Yes | No |
| 7.85 | positive | | | | | | | | | | | | | | |
| 4.52 | negative | | | | | | | | | | | | | | |

| | | | | | | | | | | | | | | | |
|---|---|---|---|---|---|---|---|---|---|---|---|---|---|---|---|
| 8.28 | positive | No | No | No | No | No | No | Yes | No | No | No | No | No | No | No |
| 8.57 | positive | No | No | No | Yes | No | Yes | Yes | Yes | No | Yes | No | No | No | No |
| 6.70 | negative | No | Yes | No | No | No | No | Yes | No | No | No | No | No | Yes | No |
| 5.49 | positive | No | No | No | No | No | No | Yes | Yes | No | No | No | No | Yes | No |
| 7.79 | positive | No | Yes | No | Yes | No | Yes | No | Yes | No | No | No | No | No | No |
| 7.36 | positive | | | | | | | | | | | | | | |
| 6.32 | positive | No | Yes | Yes | No | No | Yes | Yes | Yes | No | Yes | No | No | Yes | No |
| 7.73 | positive | No | Yes | No | Yes | No | No | Yes | No | No | No | No | No | No | No |
| 7.30 | positive | Yes | No | No | No | No | Yes | Yes | Yes | Yes | Yes | No | No | No | No |
| 5.79 | positive | No | Yes | No | Yes | No | No | No | Yes | No | No | No | No | No | Yes |
| 5.76 | positive | No | Yes | No | No | No | No | No | No | Yes | No | No | No | Yes | |
| 5.64 | positive | | | | | | | | | | | | | | |
| 6.49 | positive | No | No | No | Yes | No | No | No | Yes | Yes | No | No | No | No | No |
| 7.53 | positive | Yes | Yes | Yes | Yes | No | Yes | Yes | Yes | Yes | No | Yes | No | No | No |
| 6.82 | positive | | | | | | | | | | | | | | |
| 7.30 | positive | No | Yes | No | Yes | No | Yes | Yes | Yes | Yes | No | No | No | No | Yes |
| 5.08 | positive | No | No | No | Yes | Yes | Yes | No | Yes | No | Yes | No | No | Yes | No |
| 9.08 | positive | Yes | Yes | Yes | Yes | Yes | Yes | Yes | Yes | Yes | Yes | No | No | No | Yes |
| 7.21 | positive | No | Yes | No | Yes | No | Yes | No | Yes | Yes | No | No | No | Yes | No |
| 8.64 | positive | No | No | No | No | No | Yes | No | Yes | Yes | No | No | No | No | No |
| 9.22 | positive | No | Yes | Yes | Yes | No | Yes | Yes | Yes | No | No | No | No | No | No |
| 8.70 | positive | No | Yes | Yes | No | No | Yes | Yes | Yes | Yes | No | Yes | Yes | Yes | No |
| 4.97 | positive | No | Yes | No | No | No | No | Yes | No | Yes | No | No | | No | No |
| 8.32 | positive | Yes | No | No | Yes | No | No | No | Yes | Yes | No | No | No | No | No |
| 7.50 | positive | No | Yes | Yes | No | No | No | No | No | No | No | No | No | No | No |
| 7.63 | negative | No | Yes | No | No | No | No | No | No | No | No | No | No | No | No |
| 7.06 | positive | No | No | No | No | No | No | No | Yes | Yes | No | No | No | Yes | No |
| 7.33 | positive | No | No | No | No | No | No | No | No | Yes | No | No | No | Yes | No |
| 9.35 | positive | No | Yes | No | No | No | No | Yes | No | Yes | No | No | Yes | Yes | No |
| 8.65 | positive | No | No | No | Yes | No | No | No | No | No | Yes | No | No | Yes | No |
| 8.75 | positive | Yes | Yes | No | Yes | No | Yes | Yes | Yes | No | No | No | No | Yes | No |
| 4.72 | negative | No | No | Yes | No | No | No | Yes | Yes | No | No | No | No | No | No |
| 8.46 | positive | No | Yes | | No | No | No | No | No | Yes | No | No | | No | No |
| 9.33 | positive | No | Yes | | Yes | No | No | Yes | Yes | No | No | No | | No | No |

| | | | | | | | | | | | | | | | |
|---|---|---|---|---|---|---|---|---|---|---|---|---|---|---|---|
| 5.93 | positive | No | Yes | | No | No | No | Yes | No | No | No | No | | No | No |
| 5.99 | positive | No | No | No | No | No | No | Yes | No | No | No | No | No | No | No |
| 5.17 | positive | No | No | No | No | No | No | No | No | No | No | No | No | Yes | No |
| 9.06 | positive | No | Yes | | No | No | Yes | Yes | No | Yes | No | No | | No | No |
| 8.20 | positive | Yes | No | No | Yes | No | No | No | Yes | Yes | No | No | No | Yes | No |
| 8.93 | positive | No | Yes | Yes | No | No | Yes | Yes | Yes | Yes | No | No | No | Yes | No |
| 8.83 | positive | No | Yes | Yes | Yes | No | Yes | Yes | Yes | No | No | No | No | No | No |
| 6.78 | positive | No | No | No | Yes | No | Yes | Yes | Yes | No | No | Yes | No | Yes | No |
| 7.42 | positive | No | Yes | No | No | No | Yes | Yes | Yes | No | No | No | No | Yes | No |
| 5.66 | positive | No | Yes | Yes | Yes | No | No | No | Yes | Yes | No | No | No | No | No |
| 8.02 | positive | No | Yes | Yes | Yes | No | No | No | No | No | No | No | No | Yes | No |
| 9.52 | positive | Yes | Yes | No | Yes | No | Yes | Yes | Yes | Yes | No | No | No | Yes | No |
| 8.51 | positive | No | No | No | No | No | No | Yes | Yes | No | No | No | Yes | Yes | No |
| 8.83 | positive | Yes | No | No | No | No | No | Yes | Yes | No | No | No | Yes | No | No |
| 9.03 | positive | No | No | No | No | Yes | Yes | Yes | No | No | No | No | No | Yes | No |
| 6.06 | negative | Yes | No | No | Yes | No | Yes | No | Yes | No | No | No | Yes | No | No |
| 8.64 | positive | No | No | No | Yes | No | Yes | No | No | No | No | No | No | Yes | No |
| 5.29 | negative | No | Yes | Yes | No | No | No | No | No | No | No | No | No | Yes | No |
| 5.66 | negative | No | No | No | No | No | No | No | No | No | No | No | No | Yes | No |
| 8.88 | positive | No | Yes | | No | No | No | No | Yes | Yes | No | No | | Yes | No |
| 4.78 | positive | No | No | No | No | No | No | No | No | No | No | No | No | Yes | No |
| 8.16 | positive | Yes | Yes | | Yes | No | Yes | Yes | Yes | Yes | No | No | | No | No |
| 7.68 | positive | No | Yes | | Yes | No | Yes | Yes | Yes | No | No | Yes | | Yes | No |
| 5.98 | positive | No | No | No | Yes | No | No | Yes | Yes | No | No | No | No | No | No |
| 7.67 | positive | No | No | No | No | No | Yes | Yes | Yes | Yes | No | No | | Yes | No |
| 8.95 | positive | No | Yes | | No | No | Yes | Yes | Yes | Yes | No | No | | Yes | No |
| 8.46 | positive | Yes | Yes | | Yes | No | Yes | Yes | Yes | Yes | No | Yes | No | No | No |
| 4.43 | negative | No | Yes | No | No | No | Yes | Yes | Yes | No | No | No | No | No | No |
| 8.01 | negative | No | Yes | | No | Yes | No | No | No | No | No | No | No | No | No |
| 9.39 | positive | No | Yes | No | No | No | Yes | Yes | Yes | No | No | Yes | No | No | No |
| 6.41 | positive | No | No | No | No | No | No | Yes | No | No | Yes | Yes | | Yes | No |
| 9.10 | positive | No | No | No | Yes | No | No | Yes | No | Yes | No | No | | No | No |
| 7.57 | positive | Yes | Yes | | No | No | No | Yes | Yes | Yes | No | No | | Yes | No |
| NA | positive | No | Yes | | Yes | No | Yes | Yes | Yes | Yes | No | No | | Yes | No |

| | | | | | | | | | | | | | | | |
|---|---|---|---|---|---|---|---|---|---|---|---|---|---|---|---|
| 6.22 | positive | No | No | No | No | No | Yes | Yes | No | No | No | No | | Yes | No |
| 7.07 | positive | No | Yes | | Yes | No | No | No | Yes | No | No | No | | Yes | No |
| 4.67 | negative | No | Yes | | No | No | No | Yes | No | No | No | No | No | No | No |
| 5.02 | negative | No | Yes | | No | No | No | No | No | No | No | No | | Yes | No |
| 7.64 | positive | No | Yes | No | No | No | Yes | Yes | No | No | No | No | No | No | |
| 8.78 | positive | Yes | Yes | | Yes | No | Yes | Yes | Yes | Yes | No | No | | No | Yes |
| 5.33 | negative | | | | | | | | | | | | | | |
| 4.50 | negative | No | Yes | | No | No | No | No | No | Yes | No | No | No | No | No |
| 7.57 | positive | No | No | No | Yes | No | Yes | Yes | Yes | No | No | No | | No | No |

* $log_{10}$ RNA SARS-CoV2/ml

**(G) Table 3: Antigen-based RDT with test result, CT values and viral load for PCR positive participants in Berlin and Heidelberg**

CT-value reported here represent the E-gene genome target (similar to the target described by Corman) by increasing order. A conversion of CT-values for RT-PCR tests into viral-load was performed using quantified specific in vitro-transcribed RNA (Corman 2020 Eurosurveillance).

| Antigen RDT result | Ct value (E-Gene) | Viral load ($log_{10}$ RNA SARS-CoV2/ml) | PCR assay |
|---|---|---|---|
| **Berlin TibMolBiol** | | | |
| positive | 15·64 | 9·33 | TibMolBiol |
| positive | 16·66 | 9·03 | TibMolBiol |
| positive | 17·49 | 8·78 | TibMolBiol |
| positive | 17·61 | 8·75 | TibMolBiol |
| positive | 17·99 | 8·64 | TibMolBiol |
| positive | 18·57 | 8·46 | TibMolBiol |
| positive | 21·33 | 7·64 | TibMolBiol |
| positive | 21·57 | 7·57 | TibMolBiol |
| positive | 21·81 | 7·50 | TibMolBiol |
| negative | 26·66 | 6·06 | TibMolBiol |
| negative | 29·13 | 5·33 | TibMolBiol |
| negative | 29·27 | 5.29 | TibMolBiol |
| positive | 30·97 | 4·78 | TibMolBiol |
| negative | 31·19 | 4·72 | TibMolBiol |
| negative | 31·91 | 4·50 | TibMolBiol |
| **Berlin Roche Cobas** | | | |
| positive | 17·53 | 9·52 | Roche Cobas |
| positive | 17·97 | 9·39 | Roche Cobas |
| positive | 18·11 | 9·35 | Roche Cobas |
| positive | 18·56 | 9·22 | Roche Cobas |
| positive | 18·97 | 9·10 | Roche Cobas |
| positive | 19·09 | 9·06 | Roche Cobas |
| positive | 19·46 | 8·95 | Roche Cobas |

| | | | |
|---|---|---|---|
| positive | 19·54 | 8·93 | Roche Cobas |
| positive | 19·69 | 8·88 | Roche Cobas |
| positive | 19·86 | 8·83 | Roche Cobas |
| positive | 19·88 | 8·83 | Roche Cobas |
| positive | 20·33 | 8·70 | Roche Cobas |
| positive | 20·47 | 8·65 | Roche Cobas |
| positive | 20·52 | 8·64 | Roche Cobas |
| positive | 20·96 | 8·51 | Roche Cobas |
| positive | 21·14 | 8·46 | Roche Cobas |
| positive | 21·61 | 8·32 | Roche Cobas |
| positive | 22·01 | 8·20 | Roche Cobas |
| positive | 22·14 | 8·16 | Roche Cobas |
| positive | 22·62 | 8·02 | Roche Cobas |
| negative | 22·66 | 8·01 | Roche Cobas |
| positive | 23·77 | 7·68 | Roche Cobas |
| positive | 23·79 | 7·67 | Roche Cobas |
| negative | 23·93 | 7·63 | Roche Cobas |
| positive | 24·13 | 7·57 | Roche Cobas |
| positive | 24·64 | 7·42 | Roche Cobas |
| positive | 24·95 | 7·33 | Roche Cobas |
| positive | 25·35 | 7·21 | Roche Cobas |
| positive | 25·83 | 7·07 | Roche Cobas |
| positive | 25·85 | 7·06 | Roche Cobas |
| positive | 26·79 | 6·78 | Roche Cobas |
| positive | 28·06 | 6·41 | Roche Cobas |
| positive | 28·69 | 6·22 | Roche Cobas |
| positive | 29·46 | 5·99 | Roche Cobas |
| positive | 29·5 | 5·98 | Roche Cobas |
| positive | 29·67 | 5·93 | Roche Cobas |
| positive | 30·58 | 5·66 | Roche Cobas |
| negative | 30·59 | 5·66 | Roche Cobas |

| | | | |
|---|---|---|---|
| positive | 32·25 | 5·17 | Roche Cobas |
| negative | 32·73 | 5·02 | Roche Cobas |
| positive | 32·91 | 4·97 | Roche Cobas |
| negative | 33·92 | 4·67 | Roche Cobas |
| negative | 34·74 | 4·43 | Roche Cobas |
| **Heidelberg Seegene** | | | |
| positive | 14·3 | 9·53 | Seegene |
| positive | 15·1 | 9·30 | Seegene |
| positive | 15·8 | 9·08 | Seegene |
| positive | 15·8 | 9·08 | Seegene |
| positive | 16·0 | 9·04 | Seegene |
| positive | 16·6 | 8·86 | Seegene |
| positive | 17·0 | 8·74 | Seegene |
| positive | 17·2 | 8·68 | Seegene |
| positive | 17·2 | 8·68 | Seegene |
| positive | 17·6 | 8·57 | Seegene |
| positive | 18·0 | 8·45 | Seegene |
| positive | 18·1 | 8·41 | Seegene |
| positive | 18·4 | 8·32 | Seegene |
| positive | 18·6 | 8·29 | Seegene |
| positive | 19·5 | 8·00 | Seegene |
| positive | 19·9 | 7·88 | Seegene |
| positive | 20·0 | 7·85 | Seegene |
| positive | 20·0 | 7·85 | Seegene |
| positive | 20·2 | 7·79 | Seegene |
| positive | 20·4 | 7·73 | Seegene |
| positive | 20·8 | 7·61 | Seegene |
| positive | 20·8 | 7·61 | Seegene |
| positive | 20·8 | 7·61 | Seegene |
| positive | 20·9 | 7·53 | Seegene |
| positive | 21·1 | 7·53 | Seegene |

| | | | |
|---|---|---|---|
| positive | 21·4 | 7·45 | Seegene |
| positive | 21·7 | 7·36 | Seegene |
| positive | 21·9 | 7·30 | Seegene |
| positive | 21·9 | 7·30 | Seegene |
| positive | 22·7 | 7·04 | Seegene |
| positive | 22·7 | 7·04 | Seegene |
| positive | 22·9 | 7·00 | Seegene |
| positive | 23·5 | 6·82 | Seegene |
| negative | 23·9 | 6·70 | Seegene |
| positive | 24·4 | 6·56 | Seegene |
| positive | 24·6 | 6·49 | Seegene |
| positive | 25·1 | 6·34 | Seegene |
| positive | 25·2 | 6·32 | Seegene |
| positive | 25·2 | 6·32 | Seegene |
| positive | 27·0 | 5·79 | Seegene |
| positive | 27·1 | 5·76 | Seegene |
| positive | 27·1 | 5·76 | Seegene |
| positive | 27·5 | 5·64 | Seegene |
| positive | 28·0 | 5·49 | Seegene |
| positive | 29·4 | 5·08 | Seegene |
| negative | 31·3 | 4·52 | Seegene |
| negative | 31·8 | 4·36 | Seegene |
